# Integrated Stress Response Signatures Drive Monocyte Dysfunction in *GBA1*- and *LRRK2*-Linked Parkinson’s Disease

**DOI:** 10.1101/2025.10.08.25337448

**Authors:** Daniele Mattei, Erica Brophy, Mikaela Rosen, Oriol Narcis Majos, Aloysius Domingo, Elena Meijia, Beomjin Jang, Tarek Khashan, Mengxi Yang, Deborah Raymond, Casey Young, Jack Humphrey, Elisa Navarro, Amanda Allan, Katherine Leaver, Viktoriya Katsnelson, Alexander Chung, Benjamin Muller, Matthew Swan, Vicki L. Shanker, Mariel Pullman, Adina Wise, Roberto Ortega, Kelly Astudillo, Steven Frucht, Susan Bressman, Giulietta Riboldi, Laurie J. Ozelius, Rachel Saunders-Pullman, Towfique Raj

## Abstract

Monocytes are increasingly implicated in Parkinson’s disease (PD) pathogenesis, with idiopathic cases showing mitochondrial and lysosomal dysfunction. However, the impact of PD-associated mutations on monocytes remains unclear. To address this, we investigated transcriptomic and functional disturbances in peripheral monocytes from patients with *GBA1*- and *LRRK2*-associated PD and idiopathic PD. Transcriptomic data revealed shared and mutation-specific signatures, including those related to immune dysregulation, and consistent defects in lysosomal and mitochondrial pathways. Network and pathway analyses further uncovered downregulation in protein translation and enrichment of integrated stress response (ISR) signatures, alongside aberrant expression of genes linked to ER stress, proteostasis, mitophagy and type-I interferon signaling. These data suggest that monocyte immune dysfunction in PD may be, at least in part, a consequence of impaired proteostasis, organelle stress and maladaptive ISR activation. We further interrogated these signatures in functional assays in patient-derived macrophages, which revealed impaired mitochondrial potential, proteolysosomal dysfunction, and defective phagocytosis. Our findings define convergent molecular and functional abnormalities in genetic PD monocytes, implicating proteostasis failure and maladaptive ISR activation as upstream drivers of immune dysfunction, highlighting novel targetable pathways for therapeutic intervention.

## Introduction

Parkinson’s disease (PD) is the second most common neurodegenerative disorder and arises from a complex interplay of genetic and environmental factors^1^. Among the most prevalent genetic risk factors are variants in *GBA1*, which encodes the lysosomal enzyme glucocerebrosidase (GCase)^2–4^, and *LRRK2*, which encodes leucine-rich repeat kinase 2^5–7^. PD-associated *GBA1* mutations, such as N370S, have been proposed to reduce GBA enzymatic activity, and impair lysosomal, autophagic, and mitochondrial functions in neurons, thereby promoting α-synuclein (α-syn) misfolding and enhancing Lewy body formation.^8^ In contrast, *LRRK2* mutations, such as G2019S, result in elevated kinase activity and disruption of vesicle trafficking, lysosomal homeostasis, and mitochondrial dynamics.^8^

While most PD studies focus on neuronal dysfunction, both *GBA1* and *LRRK2* are highly expressed in myeloid cells, including brain-resident microglia and peripheral monocytes ^9,10^,. We and others have shown that PD risk variants modulate gene expression in myeloid cells^9–12^ and that monocytes from idiopathic PD (iPD) patients display mitochondrial, lysosomal/proteasomal, and immune pathway alterations^9,10,13^. Multiple studies have reported abnormal monocyte activation, impaired phagocytosis, and altered responses to α-syn in PD (reviewed in^14^). Dysfunctional monocytes may drive neurodegeneration through peripheral inflammation, altered neuroimmune signaling ^14,15^, and CNS infiltration^16–18^. Indeed, elevated pro-inflammatory cytokines and chemokines, including the monocyte chemoattractant CCL2, have been observed in both blood and cerebrospinal fluid (CSF) of PD patients, correlating with motor and neuropsychiatric symptom severity^19^. Abnormal monocyte proportions in CSF from PD patients and preclinical models further support their role in PD-related CNS pathology^17,18,20–22^. Inflammatory changes have also been observed in *GBA1*- and *LRRK2*-associated PD, including early evidence of immune pathway dysregulation, albeit with distinct features^9,23–27^. Functional studies using patient monocyte-derived macrophages and iPSC-derived microglia (iMGLs) have shown that PD-associated *GBA1* (N370S, L444P) and *LRRK2* (G2019S) mutations lead to abnormal transcriptional states, immune responses, and phagocytic activity^28–31^. We recently demonstrated that G2019S iMGLs exhibit transcriptional deregulation and enhanced myelin phagocytosis, and that monocytes from *GBA1*- and *LRRK2*-PD patients show distinct transcriptional profiles compared to iPD, particularly in immune and α-synuclein degradation pathways^9,32^.

Despite the growing evidence of altered monocyte state in PD, anti-inflammatory therapies have shown limited clinical efficacy^33^, suggesting that immune activation changes likely represent downstream effects of disrupted cellular homeostasis and underscoring the need for deeper mechanistic understanding. Monocytes from genetically defined PD cases represent a valuable model to dissect variant-specific effects and uncover convergent, targetable pathways^9^. Importantly, *GBA1* and *LRRK2* function may also be disrupted in iPD^34–37^, suggesting that insights gained from genetic forms may be broadly applicable across PD subtypes. In the present study, we profiled monocytes from *LRRK2*-PD and *GBA1*-PD cases, all from Ashkenazi Jewish (AJ) individuals, a genetically homogeneous population that minimizes confounding from population stratification. Alongside transcriptomic profiling, we performed functional assays of myeloid activity, linking genetic and transcriptional alterations to defects in mitochondrial potential, proteolysosomal function, and phagocytosis. This design enhances our ability to define disease- and mutation-specific mechanisms of monocyte dysfunction in PD and informs the development of targeted immunomodulatory therapies.

## Results

### Shared and Divergent Transcriptomic Features of Genetic and idiopathic PD

We recruited 52 individuals with PD carrying the *LRRK2* G2019S mutation, 57 *GBA1* variant carriers with PD, 124 individuals with iPD, and 41 neurologically and immunologically healthy controls from the Tom and Bonnie Strauss Movement Disorders Center and from the Bendheim Parkinson and Movement Disorders Center at Icahn School of Medicine in New York City^38,39^ (**Table 1; Supplementary** Fig. 1). All participants were of genetically confirmed AJ ancestry, with a mean age of 68.8 years at the time of blood collection (n = 274, see methods).

**Table 1:**
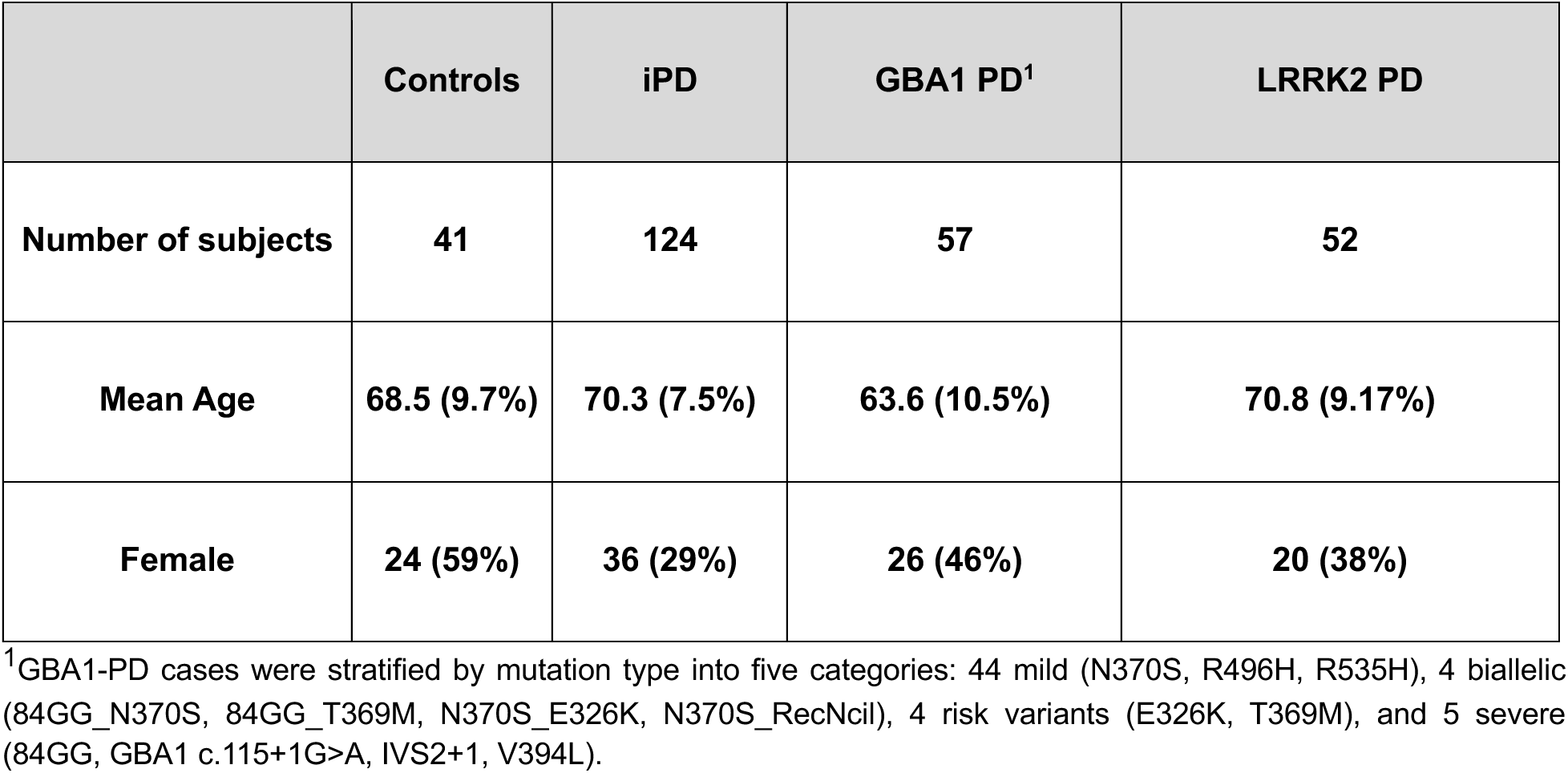
Demographic overview.

CD14⁺ monocytes were isolated from each participant for bulk RNA sequencing (RNA-seq; Supplementary Fig. 1). Following normalization and correction for known biological and technical covariates, we identified 1,595 differentially expressed genes (DEGs) in *GBA1*-PD monocytes compared to iPD monocytes (752 upregulated and 843 downregulated at FDR < 0.05; |log2-fold change (FC)| >0; **Fig. 1A**). In *LRRK2*-PD, we found 2,085 DEGs, including 1,164 upregulated and 921 downregulated genes (FDR < 0.05; **Fig. 1B**, |log2-fold change (FC)| >0; **Supplementary Table 1**). Comparison of DEG discovery in GBA1- and LRRK2-PD monocytes revealed 970 shared DEGs, with 583 upregulated and 388 downregulated genes.

**Fig. 1.**
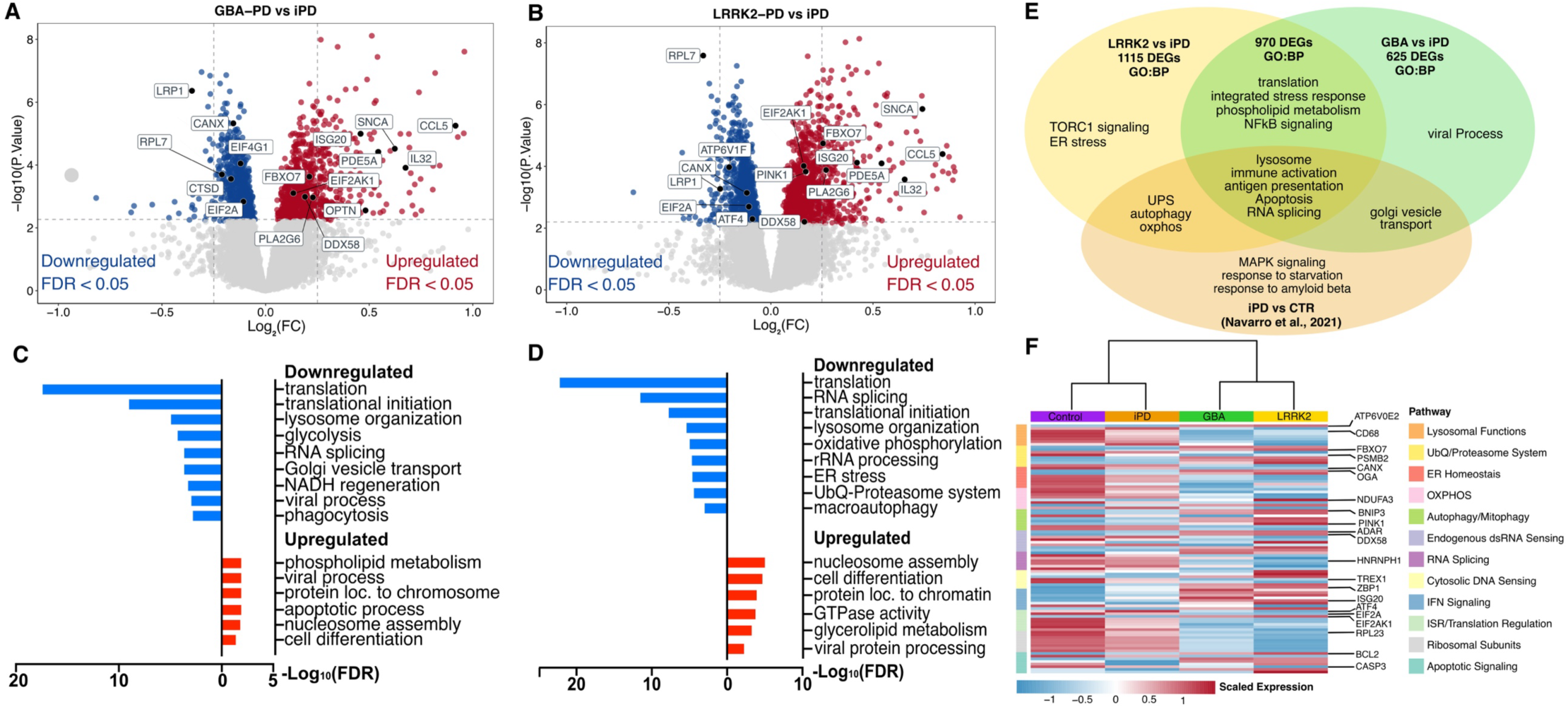
Differential gene expression analysis of idiopathic PD (iPD), GBA1-PD and LRRK2-PD monocytes. **(A-B)** Volcano plot showing the fold-change (FC) of genes (log2 scale) between *GBA1* (A) and *LRRK2* (B) PD-monocytes and iPD and their P-values significance (y-axis, −log10 scale). Differentially expressed genes (DEGs) at FDR < 0.05 are highlighted in red (upregulated genes) and blue (downregulated genes). Moderated t-statistic (two-sided) is used for statistical tests, followed by Benjamini-Hochberg adjustment. Highlighted in the volcanos are genes involved in the dysregulated biological processes associated with the DEGs. **(C-D)** Top enriched Gene Ontology (GO) biological processes (manually collapsed) for the DEGs between *GBA1*-(C) or *LRRK2*-PD (D) and iPD. The x-axis shows red for upregulated and blue for downregulated DEGs. Significance is represented in the x-axis (−log10 scale of the BH q-value). **(E)** Venn diagram showing gene ontology biological processes (BPs) for overlapping DEGs between *GBA1*- and *LRRK2*-PD monocytes, subgroup-specific DEGs (*GBA1* vs. iPD, *LRRK2* vs. iPD), and BPs enriched in iPD vs. controls (Navarro et al., 202110). **(F)** Heatmaps showing expression of DEGs (BH adjusted p < 0.05) associated with dysregulated biological processes across disease groups. Scaled expression is defined as z-score-transformed mean expression per gene, calculated by centering and standardizing corrected normalized expression data.

Gene ontology (GO) analysis revealed that upregulated DEGs in both *GBA1*-PD and *LRRK2*-PD monocytes were enriched for biological processes (BPs) such as phospholipid metabolism, viral processing, cell differentiation, and apoptotic signaling (**Fig. 1C, D**). In addition, *LRRK2*-PD upregulated pathways included mitophagy, GTPase regulation, and TOR signaling (**Fig. 1D, E, Supplementary Table 1**). Among downregulated genes, both genetic PD subtypes showed strong enrichment for protein translation, RNA splicing, lysosomal function, and type I interferon (IFN-I) signaling, and the integrated stress response (ISR) (**Fig. 1C–E, Supplementary Table 1**). *GBA1*-PD monocytes exhibited downregulation in glycolysis, phagocytosis, viral response pathways, and Golgi vesicle transport (**Fig. 1C, E, Supplementary Table 1**), while *LRRK2*-PD monocytes showed distinct downregulation of oxidative phosphorylation (OXPHOS), ER stress, ubiquitin-proteasome system (UPS), macroautophagy, as well as downregulation of translation and splicing (**Fig. 1D–E, Supplementary Table 1**).

### Transcriptional signatures of impaired clearance, ISR, and organelle stress mark GBA1- and LRRK2-PD monocytes

Among the most dysregulated processes in our pathway analysis was a consistent signature of downregulation of translation initiation and regulation genes in both GBA1- and *LRRK2*-PD monocytes, indicative of ISR activation^40^ (**Fig. 1C-E, Supplementary Table 1**). Specifically, among core ISR components, *EIF2A* was downregulated and its kinase *EIF2AK1* was upregulated in both *GBA1*- and *LRRK2*-PD monocytes, along with other core ISR genes (**Fig. 1A, B, F; Supplementary** Fig. 2B). In line with stress-induced translational repression^41,42^, we further observed broad downregulation of ribosomal protein genes (e.g., *RPL5, RPS13*) (**Supplementary** Fig. 2) and translation regulators, including reduced expression of the PD-risk gene *EIF4G1* in *GBA1*-PD monocytes^43^. The ISR effector *ATF4* was selectively downregulated in *LRRK2*-PD monocytes (**Fig. 1F, Supplementary** Fig. 2), consistent with previous findings in iPD monocytes.^44^ To investigate this more accurately, we curated gene sets for specific sub-functions pertinent to the biological processes enriched amongst the DEGs (**Fig. 1**) using GO terms and the literature (see **Supplementary Table 2**). Genes linked to lysosome biogenesis (e.g., *SORT1*) and degradative enzymes (e.g., *CTSB*) were more prominently downregulated in *GBA1*-PD monocytes, consistent with prior observations in iPSC-derived neural cells from human *GBA1* (N370S) carriers.^46^ On the other hand, lysosomal acidification genes (e.g., *ATP6V1F, ATP6V0E1*) were specifically downregulated in LRRK2-PD (**Fig. 2A, Supplementary** Fig. 2A), aligning with findings in neurons from G2019S knock-in murine models.^47^ RAB GTPases (e.g., *RAB13*) were reduced in *GBA1*-PD, while RAB effector genes (e.g., *SYTL1*) were selectively upregulated in *LRRK2*-PD. In the ubiquitin-proteasome system, genes coding for E1/E2 enzymes and deubiquitinating enzymes (DUBs) were downregulated, while Tripartite Motif (TRIM) and RING finger (RNF) E3 ligases and adaptors (e.g., PD-risk gene *FBXO7*^48^) were upregulated; proteasome subunit genes (e.g., *PSMA4*) showed the strongest downregulation in *LRRK2*-PD (**Fig. 2A, Supplementary** Fig. 2A), suggesting impaired UPS clearance as seen in neural cells with *LRRK2* or *GBA1* mutations.^49,50^ Dysregulation in genes linked to cellular clearance were accompanied by altered expression in genes relevant to ER homeostasis and stress, including downregulation in ER chaperones (e.g., *SEC63, CANX*)^51^ and upregulation in ER-stress genes such as *OGA,* a regulator of ER-stress and ISR^52^, (**Fig. 2**, **Supplementary** Fig. 2).

**Fig. 2.**
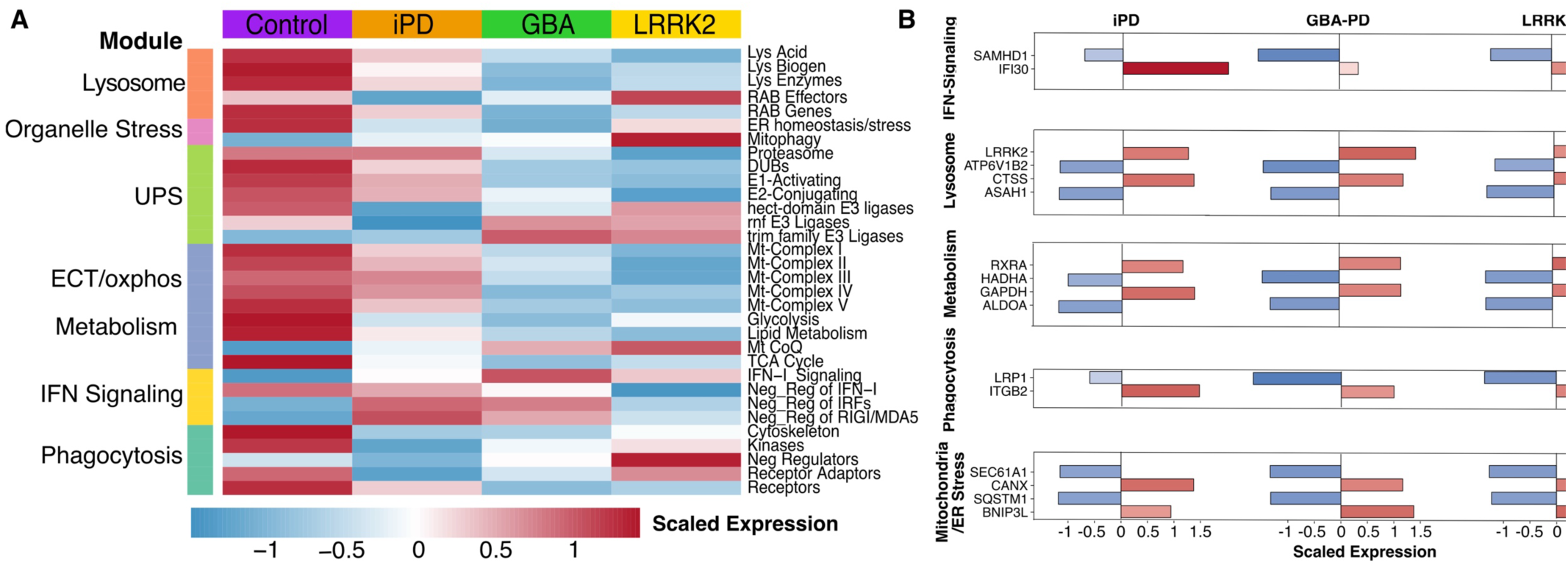
Transcriptional disruption of lysosomal, proteostatic, and metabolic pathways in GBA1-PD, LRRK2-PD, and iPD. **(A)** Heatmaps showing expression of curated gene sets representing dysregulated biological processes across the four groups. **(B)** Barplots for corrected scaled expression comparing the selected genes within pathways from (A). Scaled expression is defined as z-score-transformed mean expression per pathway (A) or gene (B), calculated by centering and standardizing corrected normalized expression data.

Mitochondrial gene-set analysis revealed widespread downregulation of mitochondrial complex I–V genes, most pronounced in *LRRK2*-PD, and upregulation of coenzyme Q (CoQ) genes, potentially compensating for electron transport chain deficits. We further observed a parallel downregulation of tricarboxylic acid (TCA)-cycle, glycolysis, and lipid metabolism genes, suggesting immunometabolic reprogramming (**Fig. 2**). Mitophagy-related genes were also elevated (e.g., *BNIP3, OPTN, MAP1LC3A*)^53^, particularly in *LRRK2-*PD monocytes, which also displayed upregulation of the PD-risk gene PINK1 (**Supplementary** Fig. 2).

Furthermore, related to the enrichment in BPs such as viral response (**Fig. 1C, D**) we observed an upregulation of type-I interferon (IFN-I) signaling genes in both groups (**Fig. 2A, Supplementary** Fig. 2A). This included upregulation of endogenous double stranded RNA (dsRNA) sensors (*RIG-I, ADAR, RNASEL*) and downregulation of dsRNA-binding proteins (*DHX9, ILF3*), consistent with prior PD blood transcriptome findings^54^ (**Supplementary** Fig. 2).

We also observed dysregulation of genes coding for cytosolic DNA sensing pathway components (*ZBP1, TREX1, SLC19A1*)^55^. Additionally, interferon-stimulated genes such as ISG15 and IFITM1, were also upregulated, **(Supplementary** Fig. 2A). Thus, by further parsing biological processes enriched in downregulated DEGs in both *GBA1*- and *LRRK2*-monocytes, we uncovered pathways known to trigger ISR and may underlie an ISR-activated state in these cells. These processes include impaired lysosomal clearance/autophagy, mitochondrial dysfunction, ER stress and activation of viral-response pathways.^40,45^

Finally, phagocytosis pathways displayed shared downregulation of phagocytic receptors, with adaptors and cytoskeleton-modulating genes upregulated in *LRRK2*-PD but downregulated in *GBA1*-PD, alongside increased expression of genes coding for negative regulators of phagocytosis and elevated phagocytic signaling kinases in both (**Fig. 2**). These transcriptional changes were accompanied by enrichment in genes linked to apoptotic signaling (e.g., *BCL2, FOXP1*) in both *GBA1* and *LRRK2*-PD groups (**Fig. 1F, Supplementary** Fig. 2A).

### Network analysis reveals mitochondrial, immune and translation modules distinguishing GBA1- and LRRK2-PD monocytes

To identify functional gene modules within gene coexpression networks, we applied Weighted Gene Co-expression Network Analysis (WGCNA, **Supplementary** Fig. 4) to monocyte RNASeq data from iPD, *GBA1*-PD, *LRRK2*-PD, and controls. This analysis yielded 53 modules, 13 of which (M1–M13) had eigengenes significantly associated with Parkinson’s disease diagnosis. (**Fig. 3A-I**). These modules were enriched for protein translation (M1, M3-M5), proteasomal functions (M1), stress response (M12) and mitochondrial functions, (M1, M3, M4, M7 and M10) (**Fig. 3A-I, Supplementary Table 3**). Modules M11-M13 were enriched for genes linked to cell migration, interferon signaling, immune response and cholesterol metabolism.

**Fig. 3.**
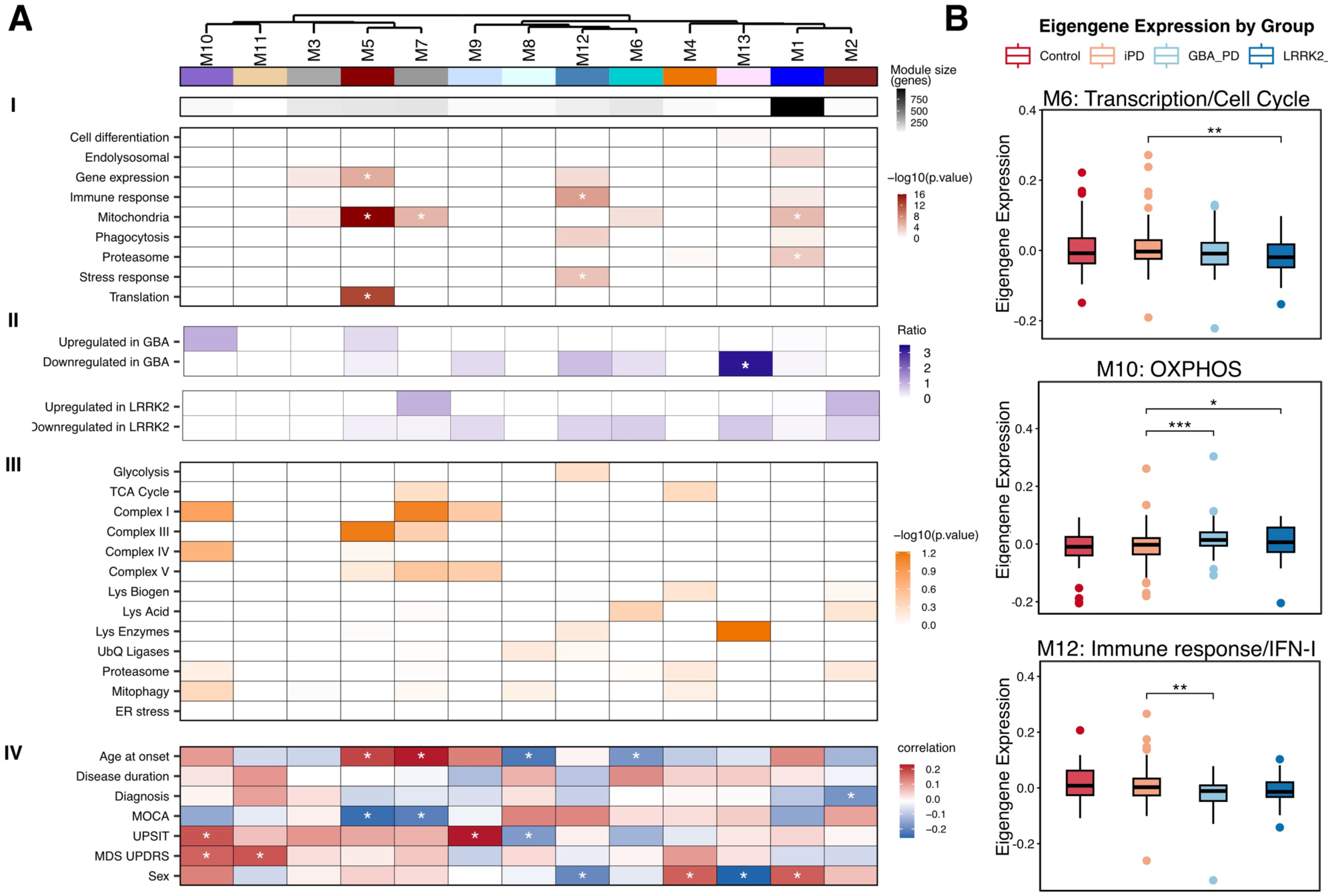
Weighted gene co-expression analysis in PD monocytes. **(A)** Weighted gene co-expression network analysis (WGCNA) of 274 monocyte samples (Control, iPD, GBA1-PD, LRRK2-PD) identified 13 significant gene modules. Modules are shown as a hierarchical clustering based on module eigengene (ME) correlation (top). Panels I–IV display associations between modules and: **(I)** GO biological process enrichment (manually collapsed); **(II)** Gene ratio refers to the enrichment of WGCNA module genes within the up- or down-regulated differentially expressed genes for GBA-PD and LRRK2-PD monocytes compared to iPD monocytes, (Fisher’s exact test, adjusted p < 0.05); (**III)** curated gene sets for key biological processes; and **(IV)** Spearman correlations with clinical traits. Asterisks indicate q < 0.05. P-values: (I) hypergeometric test; (II, III) one-sided Fisher’s exact test with BH adjustment; **(IV)** Spearman rank correlatexpression for selected modules: M6, M10 and M12. Boxplots indicate median, quartiles, and whiskers (1.5× IQR). Eigengene expression was compared via Wilcoxon sum-rank test, nominal p-value thresholds: *p ≤ 0.05, **p ≤ 0.01, *p ≤ 0.001. MoCA: Montreal Cognitive Assessment; UPSIT: University of Pennsylvania Smell Identification Test; MDS-UPDRS: Movement Disorder Society Unified Parkinson’s Disease Rating Scale. TCA: tricarboxylic acid cycle; Lys: lysosomal; Biogen: biogenesis; Acid: acidification; UbQ: ubiquitin; ER: endoplasmic reticulum.

We next assessed how these modules relate to gene expression changes in *GBA1*-PD and *LRRK2*-PD by testing the enrichment of up- and downregulated DEGs (**Fig. 3A-II**). Module M13 genes were significantly enriched with downregulated DEGs of *GBA1*-PD monocytes. This module contained genes related to glial activation, ER metabolic pathways and cholesterol metabolism. The distribution of other modules across the DEG sets exhibited consistent patterns with our gene ontology analyses findings. Module M2, which contained genes relevant to autophagy and proteasomal functions, were enriched with up- and downregulated DEGs in *LRRK2*-PD, while M6 (transcription and cell cycle regulation) contained *LRRK2*-PD downregulated DEGs. Modules related to oxidative phosphorylation (M7, M9, M10) were enriched with up- and downregulated DEGs in *GBA1*- and *LRRK2*-PD. Module M12 genes, enriched for immune activation and IFN-I signaling functions, included downregulated DEGs in both groups. To further refine the modules’ biological relevance, we annotated gene modules with curated gene sets for metabolism, lysosomal, and mitochondrial pathways used in Fig. 2, (**Fig. 3A-III, Supplementary Table 2**). Mitochondrial complex I/III and lysosomal acidification genes were overrepresented in modules strongly dysregulated in *LRRK2*-PD, whereas lysosomal enzyme genes and mitochondrial complex I/IV components were prominent in *GBA1*-PD-associated modules.

We used module eigengenes to associate coexpression modules to clinical features. Older age of onset correlated positively with M5 and M7 expression (translation, mitochondrial complex I/III) and correlated negatively with Montreal Cognitive Assessment (MoCA) scores (**Fig. 3A-IV**). Conversely, younger onset was positively associated with the eigengenes of M6 (cell cycle, transcriptional regulation) and M8 (endosomal transport). PD diagnosis was negatively associated with the expression of module M2 (signal transduction, autophagy), while University of Pennsylvania Smell Identification Test (UPSIT) and Movement Disorder Society Unified Parkinson’s Disease Rating Scale (MDS-UPDRS, parts I-III) scores correlated with eigengenes of mitochondria and cell migration-related modules M10 and M11 (**Fig. 3A-IV**). Notably, sex-dependent expression differences were observed in M1, M4, M12, and M13, consistent with our previous findings of mutation specific sex-differences in risk for PD.^56^ Finally, comparison of module eigengenes between groups revealed PD-subtype specific differences (**Fig. 3B, Supplementary** Fig. 5). The eigengene of module M6, which contains genes relevant to transcriptional regulation and cell cycle, was decreased in *LRRK2*-PD monocytes relative to iPD, while the eigengene of module M10, enriched for OXPHOS genes was increased in both genetic PD-subtypes as compared to iPD. Module M12, linked to immune response and IFN-I signaling, was selectively downregulated in *GBA1*-PD monocytes relative to iPD (**Fig. 3B, Supplementary** Fig. 5).

### Functional analysis of mitochondrial, phagocytic, and proteolysosomal functions in PD monocyte-derived macrophages

To validate key transcriptional findings, we assessed mitochondrial, proteolysosomal, and phagocytic functions in monocyte-derived macrophages (MDMs) from a subset of donors. Assays were quantified by live-cell fluorescence imaging over 20–24 hours. Fluorescence values were adjusted for technical and biological covariates (see Methods) and modeled using non-linear mixed-effects regression (NLME) to examine fluorescence dynamics over time. Area under the curve (AUC) was calculated for each donor and compared across groups.

Given the transcriptional dysregulation in OXPHOS genes, we examined mitochondrial membrane potential using the potentiometric dye tetramethyrhodamine, methyl ester (TMRM^57^). NLME modeling of residual TMRM fluorescence revealed a significant baseline group effect for *LRRK2*-PD compared to control (β = –8.47×10⁶ ± 2.66×10⁶, t(61) = –3.18, p = 0.0023), indicating a lower starting TMRM signal (**Fig. 4A**). Additionally, there was a significant group-by-time interaction for *LRRK2* (β = 2.97×10⁵ ± 9.31×10⁴, t(1231) = 3.19, p = 0.0015), and a trend for *GBA1* (p = 0.071), consistent with a steeper decline in mitochondrial membrane potential over time. Analysis of the areas under the curve (AUC) showed significant overall group differences (Kruskal–Wallis test: H(3) = 9.71, p = 0.021). Post-hoc analysis confirmed reduced TMRM AUC in *LRRK2*-PD versus control (FDR = 0.038) and versus iPD (FDR = 0.032). Next, we compared the mitochondrial response to the chemotactic damage-associated signal adenosine diphosphate (ADP) between groups (**Fig. 4B**). The *LRRK2*-PD group showed a significantly lower overall TMRM signal compared to controls (β = –6.53×10⁶ ± 3.24×10⁶, t(48) = –2.01, p = 0.05), as well as a significant group × time interaction (β = 8.89×10⁴ ± 3.73×10⁴, t(1244) = 2.39, p = 0.017), indicating a decreased mitochondrial response to ADP. AUC analysis did not reveal overall significant differences in ADP response.

**Fig. 4.**
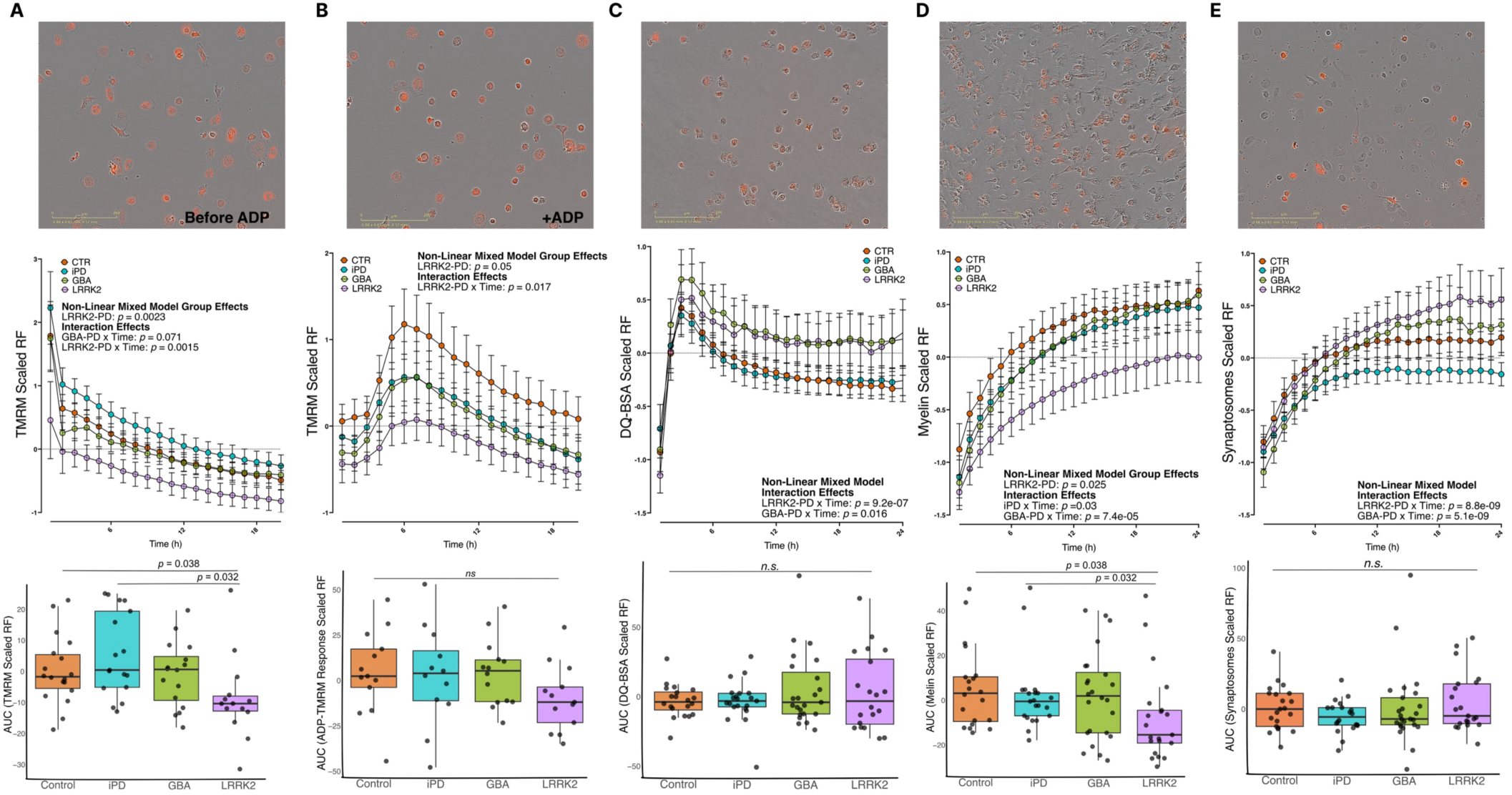
Mutation-specific functional changes in mitochondrial, lysosomal and phagocytic functions. Live-cell imaging assays were performed on monocyte-derived macrophages (MDMs) from controls (CTR), diopathic PD (iPD), GBA1-PD, and LRRK2-PD patients. Upper panels: representative pictures taken via the IncuCyte S3 apparatus during live imaging. Middle panels: Curves for the scaled residual fluorescence (RF, Y-axis) over time (Xaxis, hours) for each functional assay. Lower panels: Boxplots comparing the areas under the curve (AUC) for each functional assay between groups. (A, B) Mitochondrial membrane potential measured via the fluorescent tetramethylrhodamine (TMRM baseline (A; n = 18 CTR, 17 iPD, 16 GBA1-PD, 14 LRRK2-PD) and following ADP stimulation (100 uM) (B; n = 13 CTR, 11 iPD, 14 GBA-PD, 12 LRRK2-PD). (C) Dye quenched-bovine serum albumin-Red (DQ-BSA) fluorescence was used to measure lysosomal proteolytic activity over 24h, (n = 20 CTR, 21 iPD, 23 GBA1-PD, 20 LRRK2-PD). (D) pHRodo-tagged myelin debris (n = 20 CTR, 21 iPD, 24 GBA1-PD, 21 LRRK2-PD) and (E) pHRodo-tagged synaptosomes (n = 21 CTR, 20 iPD, 25 GBA1-PD, 21 LRRK2-PD) were used as CNS-relevant substrates to assess phagocytic function in response to neurodegeneration-related stimuli. Fluorescence values were adjusted for technical and biological covariates and analyzed as residualized fluorescence and plotted as scaled residual fluorescence. Curve dynamics were modeled using non-linear mixed-effects regression with group, time, and their interaction as fixed effects. Area under the curve (AUC) was calculated for each sample and compared across groups using Kruskal-Wallis tests followed by FDR-corrected pairwise Wilcoxon rank-sum tests. Curve error bars show mean ± SEM.

We next assessed MDMs proteolysosomal functions via dye quenched-bovine serum albumin-Red (DQ-BSA). The latter is internalized via endocytosis and, upon fusion with lysosomes, is degraded by lysosomal hydrolases such as CTSB^58^. This process releases a red fluorescent signal proportional to both substrate accumulation and proteolytic efficiency. A significant group by time interaction was detected for the *LRRK2*-PD group (β = 9.50×10⁵ ± 1.93×10⁵, t(1928) = 4.92, p = 9.2×10⁻⁷) and the *GBA1*-PD group (β = 4.49×10⁵ ± 1.87×10⁵, t(1928) = 2.40, p = 0.016), indicating a steeper trajectory of DQ-BSA signal accumulation over time, consistent with altered lysosomal processing efficiency (**Fig. 4C**).

We then used pHRodo-tagged myelin debris (**Fig. 4D**) and synaptosomes (**Fig. 4E**) as CNS-associated phagocytic stimuli relevant to neurodegeneration to compare MDMs phagocytosis between groups. We deliberately did not use pathological α-synuclein as a phagocytic stimulus, given its well-documented ability to induce strong inflammatory responses in myeloid cells, which can confound the interpretation of intrinsic phagocytic capacity.^59,60^ Instead, we selected more neutral substrates to assess phagocytosis in a controlled and physiologically relevant manner. pHRodo is a pH-sensitive dye that progressively emits increasing fluorescence as the phagosome fuses with lysosomes^61^. Curve analysis revealed a significant main group effect for *LRRK2*-PD (β = –5.72×10⁶ ± 2.50×10⁶, t(82) = –2.29, p = 0.0249), indicating overall reduced phagocytic capacity. Time-dependent group differences were detected for *GBA1*-PD (group × time interaction: β = 1.58×10⁵ ± 3.97×10⁴, t(1974) = 3.97, p = 7.4×10⁻⁵) and iPD (β = 8.92×10⁴ ± 4.10×10⁴, t(1974) = 2.18, p = 0.0295), suggesting altered myelin phagocytic dynamics in GBA1 and iPD groups. AUC comparisons showed significant overall group differences (Kruskal–Wallis H(3) = 9.52, p = 0.023). Post-hoc analysis confirmed a reduced cumulative myelin phagocytosis in *LRRK2*-PD versus control (FDR = 0.03) and versus iPD (FDR = 0.03). Finally, pHRodo-synaptosome phagocytosis dynamics revealed significant group-by-time interaction effects for both *GBA1-*PD (β = 4.87×10⁵ ± 8.30×10⁴, t(1997) = 5.87, p = 5.13×10⁻⁹) and LRRK2-PD (β = 5.00×10⁵ ± 8.65×10⁴, t(1997) = 5.78, p = 8.82×10⁻⁹), indicating enhanced synaptosome phagocytosis dynamics over time in *GBA1*- and *LRRK2*-PD monocytes.

## Discussion

In this study, we provide a comprehensive transcriptomic and functional characterization of peripheral monocytes in iPD and genetic PD (*GBA1*- and *LRRK2*-PD) within a genetically homogeneous AJ cohort. By integrating transcriptomic and functional analyses, we identify shared and mutation-specific disruptions in lysosomal, mitochondrial, and proteostasis pathways converging on ISR activation and impaired innate immune function.

Across genetic PD subtypes, we observed downregulation of the translation machinery, lysosomal function, proteostasis pathways, and interferon signaling, accompanied by ISR signatures. These transcriptional alterations mirror prior findings in iPD neurons, monocytes and microglia^10,40,62^, underscoring shared mechanisms linking impaired protein clearance, disrupted metabolic homeostasis, and immune dysfunction in PD. Over 970 DEGs were shared between *GBA1*- and *LRRK2*-PD monocytes, converging on downregulated translation, lysosomal dysfunction, ISR, and immune activation. We found an overlap with our previous findings in iPD monocytes relative to controls,^10^ which displayed deregulation in genes linked to oxidative phosphorylation, lysosomal functions, and the UPS, pointing to common impairments in clearance and proteostasis across PD subtypes, shaped by both genetic and idiopathic PD.

We also observed distinct molecular signatures in *GBA1*- and *LRRK2*-PD monocytes. *GBA1*-PD was marked by deficits in lysosome biogenesis and enzyme genes, phagocytic receptors, and viral response pathways, aligning with impaired proteolysosomal degradation and altered phagocytosis in MDM assays. In contrast, *LRRK2*-PD showed pronounced downregulation of OXPHOS, proteasome components, and ER-stress regulators, coupled with mitophagy gene upregulation and reduced mitochondrial membrane potential, suggesting a compensatory but insufficient response to bioenergetic stress. These findings indicate that *GBA1*-PD, as anticipated, primarily affects lysosomal enzyme-driven degradation, while *LRRK2*-PD additionally impairs mitochondrial metabolism and proteostasis, aligning with prior reports of hyperactive *LRRK2* kinase-driven vesicular and mitochondrial dysfunction. While these distinct transcriptional profiles characterize the two genetic PD subtypes, the final common pathway in monocytes appears to be a sustained ISR, leading to monocyte activation, and contributing to immune cell dysfunction in PD regardless of the genetic cause.

ISR suppresses global protein synthesis via phosphorylation of the eukaryotic translation initiation factor eIF2α in response to impaired proteostasis, ER stress and mitochondrial dysfunctions^40^. We found marked downregulation of translation-related genes, alongside reduced EIF2A expression and increased levels of the kinase EIF2AK1, a key ISR regulator implicated in α-synuclein-induced stress.^41,42^ While the ISR is increasingly investigated in the context of neurodegeneration, where the aforementioned triggers have been implicated in neuronal vulnerability and death^40^, our data suggest similar mechanisms may underlie monocyte dysfunction in genetic PD.

We hypothesize that impaired lysosomal degradation and disrupted UPS contribute to defective proteostasis, contributing to ER stress and mitochondrial dysfunction in *GBA1*- and *LRRK2*-PD monocytes (**Fig. 5**). These upstream alterations likely converge to trigger ISR activation. Furthermore, we observed broad dysregulation of genes involved in OXPHOS and mitophagy, paired with reduced mitochondrial membrane potential, indicative of accumulated damaged mitochondria. Damaged mitochondria can release mitochondrial RNA (mtRNA), which can form double-stranded RNA (dsRNA) structures^63^, and mitochondrial DNA (mtDNA) into the cytosol^64^. These nucleic acids act as viral mimics, engaging cytosolic sensors and promoting IFN-I signaling, which can perpetuate ISR activation^65^. Indeed, we observed enrichment of viral and IFN-related pathways, with upregulation of dsRNA and cytosolic DNA sensors (e.g., ADAR, RIG-I, ZBP1) and interferon-stimulated genes. Moreover, consistent with our findings and the growing literature on altered monocyte activation in PD, chronic ISR in myeloid cells has been associated with altered inflammatory responses, metabolic rewiring, and apoptotic signaling^45,66,67^.

ISR signatures have also been identified in microglia from Alzheimer’s disease (AD) brains and mouse models^68^, and more recently in iPD microglia^62^, supporting conserved stress responses across neurodegenerative diseases and myeloid populations. However, the latter study does not provide mechanistic insights into potential upstream causes of the microglial ISR in PD. Our data point to impaired clearance and proteostasis as candidate upstream drivers. Importantly, similar transcriptional programs have been reported in whole blood and patient-derived cells from idiopathic and *LRRK2*-PD cases^69,70^, further suggesting that translational repression may be shared features of PD. ISR-related signatures have also been reported in peripheral blood of prodromal and early-stage PD patients, and in postmortem substantia nigra from early and late PD stages^54,71,72^, underscoring its relevance across disease progression and compartments.

The observation of ISR signatures across brain-resident microglia and peripheral monocytes raises important questions about their origin and triggers. Microglia are tissue-resident and directly exposed to toxic α-synuclein, a well-established inducer of lysosomal, proteasomal, and ISR dysregulation.^40,73^ In contrast, classical monocytes are short-lived (1-3 days^74^) and not tissue-resident, making the presence of comparable signatures in monocytes from both idiopathic and genetic PD particularly intriguing. Our results suggest that cell-autonomous defects in clearance and proteostasis may, at least in part, be driven by PD-linked mutations (e.g., *GBA1, LRRK2* in the present study) or, in iPD, by the intersection of common risk variants^10^ and environmental factors. Importantly, several studies demonstrate that *ex vivo* exposure of monocytes to pathogenic α-synuclein triggers pro-inflammatory activation and impaired autophagy.^59,75,76^ Thus, future research should aim at dissecting the potential contribution of peripheral blood α-synuclein to monocyte dysfunctions in PD patients.

In summary, our study implicates active ISR in monocytes accompanied by functional impairments in mitochondrial potential, lysosomal degradation, and phagocytosis as contributory to the molecular alterations and innate immune dysfunction in genetic forms of PD (**Fig. 5**). This study is limited by its cross-sectional design, precluding causal inference regarding whether monocyte dysfunction drives PD pathology or reflects a response to it or other peripheral triggers. Medications (e.g., L-DOPA, MAO-B inhibitors, anti-inflammatory drugs) and acute infections can also shape monocyte states and may confound interpretation. While we used a genetically homogeneous cohort to reduce confounding, replication in diverse populations is necessary. Functional assays were limited to ex vivo MDM models, which may not fully recapitulate in vivo immune states. Given the consistency in transcriptional signatures across PD subtypes, future work should integrate longitudinal sampling, single-cell resolution, and *in vivo* validation to dissect ISR dynamics and triggers with temporal resolution to inform both biomarker discovery and therapeutic development.

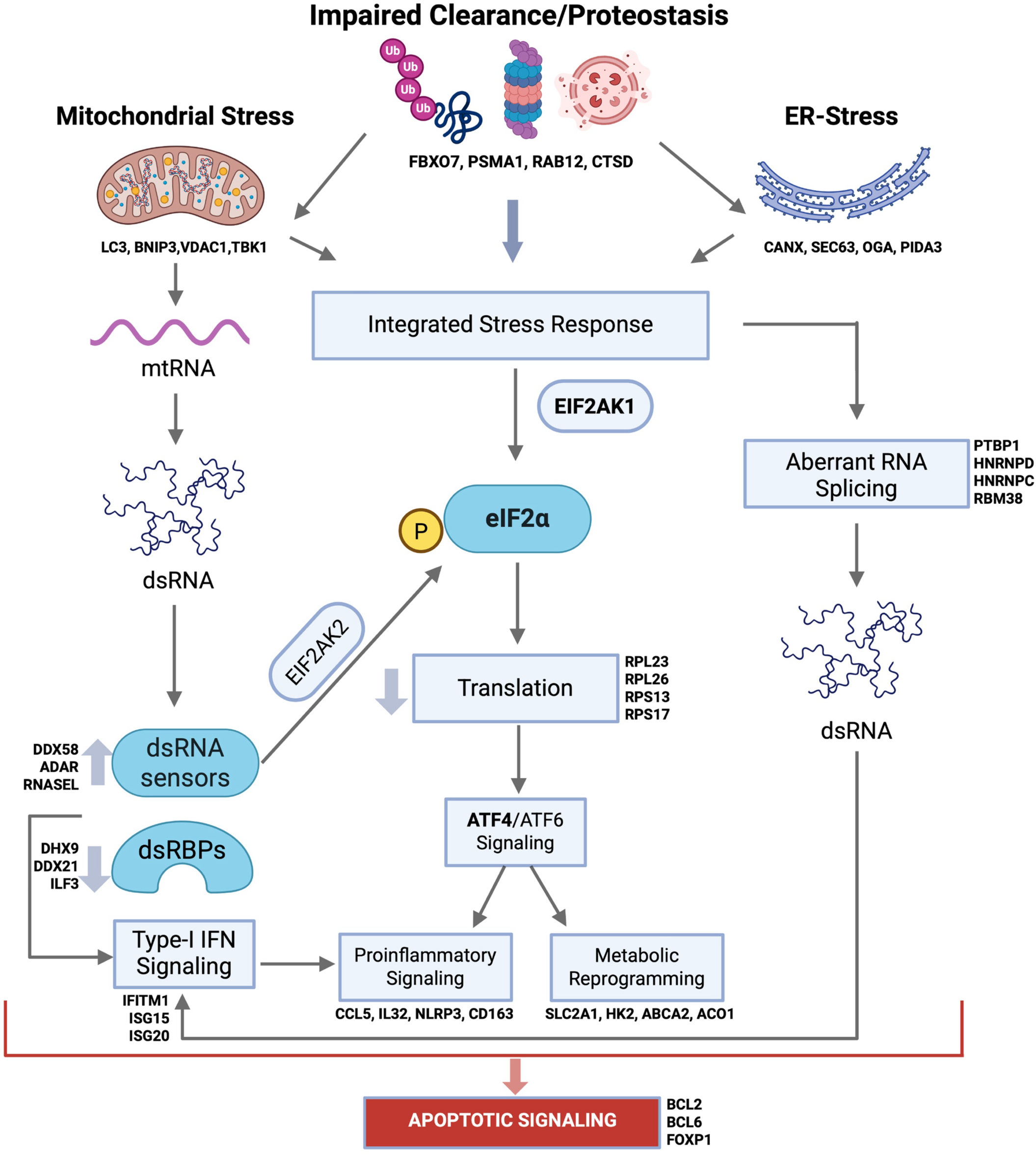
Defective clearance and organelle stress drive maladaptive monocyte immune states in GBA1- and LRRK2-PD. Schematic representation of the cellular and molecular mechanisms driving altered immune state in GBA1- and LRRK2-PD monocytes. Our transcriptional and functional profiling reveals lysosomal and proteasomal dysfunction that converge into impaired proteostasis, autophagy flux and subsequent endoplasmic reticulum (ER) and mitochondrial stress. Impaired clearance, proteostasis and organelle stress can trigger a state of integrated stress response (ISR) and induce a global decrease in translation. Additionally, leakage of nucleic acids from damaged mitochondria (e.g., mtRNA) along with aberrant RNA-splicing can increase the endogenous double stranded RNA (dsRNA)-burden, which can trigger interferon signaling through cytosolic sensors. Chronic ISR ultimately results in altered immune reactivity, metabolic reprogramming and induction of apoptotic signaling cascades. In bold are examples of genes relevant to each biological process that were significantly differentially expressed in either or both GBA1- and LRRK2-PD monocytes relative to iPD in our transcriptomic analysis. Figure generated with Biorender.

## Methods

### Clinical centers and recruitment strategies

Subjects were enrolled from the Bonnie and Tom Strauss Movement Disorders Center and the New York Movement Disorder (NYMD) cohort at the Bendheim Parkinson and Movement Disorders Center (BPMD) at the Icahn School of Medicine at Mount Sinai. Additional subjects were also included from other PD and aging cohorts including Marlene and Paolo Fresco Institute for Parkinson’s and Movement Disorders at NYU Langone Health (New York); the Alzheimer’s Disease Research Center (ADRC) at Mount Sinai; and the Center for Cognitive Health (CCH) at Mount Sinai Hospital (New York). Each institution’s Institutional Review Board (IRB) approved the study protocol, recruitment procedures, and collection of data and biospecimens. Written informed consent was obtained from all participants prior to enrollment. PD diagnoses were confirmed by movement disorder specialists according to the United Kingdom Parkinson’s Disease Society Brain Bank Clinical Diagnostic Criteria^77^, but included those with family history. Healthy controls (CTRL) were individuals with no personal history of neurological disease and no first- or second-degree relatives affected by a neurological condition.

### Blood Collection and PBMC, monocyte Isolation and RNA extraction

Blood samples were collected fresh during the morning of each research visit to minimize variability in sample composition and cell activation. Samples were drawn into Vacutainer tubes containing acid citrate dextrose (ACD) (BD Biosciences) and processed within 3–4 hours. DNA was extracted from 0.5 ml of whole blood using the QIAamp DNA Blood Midi Kit (Qiagen) following the manufacturer’s protocol, and DNA quality and concentration were assessed using a Nanodrop spectrophotometer. Blood samples were processed as previously described^10^. Briefly, processing consisted in isolation of peripheral blood mononuclear cells (PBMC) and subsequent CD14+ monocytes purification. For PBMC isolation, SepMate tubes (StemCell Technologies) were used. After dilution in 2-fold PBS (Gibco) tubes were filled with 15 ml of Ficoll-Plaque PLUS (GE Healthcare) and centrifuged at 1200g for 10 mins, followed by wash with PBS. 5 million PBMCs were immediately used for automated monocyte isolation through the AutoMacs sorter (Miltenyi) with human CD14+ magnetic beads (Miletenyi), according to manufacturer’s instructions. Sorted monocytes were stored at −80°C in RLT buffer (Qiagen) + 1% 2-Mercaptoethanol (Sigma Aldrich). Isolated monocytes stored in RLT buffer were first thawed on ice. RNA was isolated with the RNeasy Mini kit (Qiagen) according to manufacturer’s instructions, including the DNase I optional step. RNA was then stored at −80°C until library preparation. RNA integrity number (RIN) was assessed with TapeStation using Agilent RNA ScreenTape System (Agilent Technologies). RNA concentration was obtained with Qubit. The remaining PBMCs were cryopreserved in 90% FBS (Germini) + 10% dimethyl sulfoxide (DMSO, Sigma Aldrich) at a concentration of 10 million cells/ml in Nalgene cryogenic vials (ThermoScientific).

### Monocyte isolation

Monocytes were isolated from cryopreserved PBMCs by Magnetic-Activated Cell Sorting (MACS; Miltenyi) using human CD14 antibody-conjugated magnetic microbeads (Miltenyi), following protocols previously established.^10^ Sorted monocytes were plated at a density of 25,000 cells per well in poly-D-lysine-coated 96-well plates in DMEM/F12 medium (Gibco) supplemented with 10% fetal bovine serum (FBS), Glutamax, and 1% penicillin-streptomycin. Differentiation into macrophages was induced by adding human recombinant M-CSF (50 ng/mL; Peprotech), with media changes performed every third day. Functional maturity was reached at day in vitro 7 (DIV7), all assays were thus performed at DIV7. Functional assays were conducted in triplicate using the IncuCyte S3 live-cell imaging system (Sartorius). Bright-field and fluorescent images were acquired every hour and analyzed using the Incucyte Software (v2024b). Integrated fluorescence intensity was normalized to cell confluency and used for downstream quantification. All assays were performed with the experimenter blinded to the group identity of the samples.

### Functional assays

#### Mitochondrial membrane potential

In order to collect longitudinal data on the mitochondrial membrane potential, cells were stained with 25 nM tetramethylrhodamine, methyl ester (TMRM, ThermoFisher) and left in the media, as previously described^57^. At this concentration TMRM does not interfere with mitochondrial functions and its concentration within the mitochondrial matrix is low enough to ensure fluorescence intensity is proportional to the membrane potential, without self-quenching artifacts.^57^ Following 20h baseline measurements, 100 μM ADP (MilliporeSigma) were added to test the mitochondrial response to a chemotactic, damage-associated stimulus.^78^

#### Proteolysosomal functions

Dye quenched bovine serum albumin-Red (DQ-BSA-Red, ThermoFisher), was employed to measure lysosomal proteolytic functions. Cells were stained at 10 μg/ml for 30 minutes at 37°C according to manufacturer’s instructions. Following incubation, cells were washed x2 with warm cell culture media. This way, the fluorescence signal post-wash is proportional to the cellular DQ-BSA accumulation and degradation efficiency.

#### Myelin and synaptosome preparation

In order to create biologically representative samples, minimizing single-donor biases in myelin and synaptosome composition, the latter were extracted and pooled from frozen postmortem occipital cortex tissue of 8 male non-neurological control donors (mean age: 71 ± 14 years). Myelin was extracted via percoll gradient centrifugation following the protocol described in ^79^. Briefly, brain tissue was thawed in ice-cold PBS (Gibco) + protease inhibitors (Roche cOmplete protease inhibitor cocktail) and dissociated in a dounce homogenizer (Millipore Sigma). Homogenized brain was pelleted, resuspended in 30% percoll in PBS, overlaid with PBS and centrifuged at 3000 xg, 10 minutes at 4°C to separate myelin from cellular and tissue debris. Myelin was collected between the layers, washed x2 in ultra-pure distilled water to osmotically shock any remaining cell and then washed 2x in PBS. Synaptosomes were purified from the same tissue samples using the Syn-PER™ Synaptic Protein Extraction Reagent (Thermofisher) + protease inhibitors, following manufacturer’s instructions. Stocks of myelin and synaptosomes were prepared in PBS with a final volume adjusted to obtain a final protein concentration of 1μg/μl. Endotoxin levels were measured using a commercial kit following manufacturer instructions (GenScript). Myelin and synaptosomes fragments were conjugated with 1:100 pHRodo red dye (Thermofisher) according to the manufacturer’s protocol and stored at −80°C.

#### Phagocytosis assay

2.5μg of pHRodo-tagged myelin and synaptosomes respectively were added to 150μl media/well and cells were imaged hourly over 24h.

#### Statistical analysis

Integrated red fluorescence data (normalized/cell confluency) was analyzed in R (v4.5.0). For time-course functional assays, fluorescence intensity values were first adjusted for biological covariates, including subjects’ age at draw, sex, ancestry, clinic, and technical covariates, such as PBMC cryopreservation time, assay date and batch, by fitting a linear model and extracting the residuals. Residualized fluorescence values were then analyzed using a nonlinear mixed-effects (NLME) regression model with experimental group, time, and their interaction as fixed effects, and sample ID as a random intercept. Residual plots were inspected to assess model assumptions. Curves were plotted as scaled residual fluorescence intensity. Area under the curve (AUC) for each sample was computed using the trapezoidal method. Group-level differences in AUC were assessed using a Kruskal-Wallis test, followed by pairwise Wilcoxon rank-sum tests with FDR correction.

#### RNA sequencing

RNA-seq libraries were prepared using the TruSeq Stranded Total RNA Sample Preparation Kit with the Low Sample (LS) protocol (Illumina) following the manufacturer’s instructions. RNA libraries were prepared by a commercial service (Azenta Inc., formerly Genewiz Inc.) using a standard RNA-seq protocol. For all samples, ribosomal RNA was removed using a ribo-depletion strategy. Sequencing was performed at Azenta Inc. on an Illumina HiSeq 4000 platform, generating 150-bp paired-end reads with an average depth of 60 million reads per sample. Libraries were processed in four independent sequencing batches.

#### SNP and GBA1/LRRK2 Genotyping

DNA samples were genotyped using the Illumina Infinium Global Screening Array (GSA), which includes a genome-wide backbone of 642,824 common variants and ∼60,000 custom disease-related SNPs. Targeted genotyping for common *GBA1* and *LRRK2* mutations was performed at Dr. William Nichols’ laboratory (Cincinnati Children’s Hospital). For *LRRK2*, the G2019S variant was screened. For *GBA1*, 11 variants that are more frequent in the Ashkenazi Jewish population were analyzed: IVS2+1, 84GG, E326K, T369M, N370S, V394L, D409G, L444P, A456P, R496H, and RecNcil. Mutation frequencies were calculated for the entire cohort as well as for manifesting and non-manifesting carriers.

#### Genotyping QC and Ancestry Analysis

Quality control (QC) filters included minor allele frequency (MAF) > 5%, SNP and sample call rates > 95%, and Hardy–Weinberg equilibrium (HWE) *P* > 1 × 10⁻⁶. Duplicate or related samples were identified and removed using pairwise identity-by-descent (IBD) estimation in PLINK (PI_HAT = 0.99–1).

Genetic ancestry was assessed by principal component analysis (PCA) and multidimensional scaling (MDS), comparing the study cohort with Phase 3 reference samples from the 1000 Genomes Project^80^. For AJ ancestry, analyses were repeated using a custom AJ reference panel^81^. Non-AJ samples were excluded using Somalier v0.2.12^82^, with reference space derived from AMP-PD genotypes, following the approach of Iwaki et al.^83^.

#### Data processing and normalization of RNA-seq

For each cohort, gene-level expression was quantified using the GENCODE v30 genome reference and the RAPiD RNA-seq pipeline^84^. The RAPiD-nf pipeline, built on Mount Sinai’s APOLLO framework and implemented in Nextflow, processed paired-end FASTQ files through automated alignment, quantification, and quality control (QC)^85^. QC metrics were generated using Picard (v2.20)^86^ and FASTQC (v0.11.8)^87^. Trimming and alignment were performed with Trimmomatic (v0.36)^88^ and STAR (v2.7.2a)^89^, respectively. Gene and isoform abundance were estimated using RSEM (v1.3.1)^90^, and mapped reads were annotated to genomic features using featureCounts (v1.6.3) to generate read summarizations^91^.

#### Differential gene expression analysis

Differential gene expression analysis of monocytes was performed using edgeR (v4.0.16), limma (v3.58.1)^92^, and SVA (v3.5)^93^. Genes with a median TPM < 1 were excluded. Surrogate Variable Analysis (SVA) was applied to estimate and remove unknown sources of variation, while preserving mutation and disease status (**Supplementary** Fig. 3); 11 significant surrogate variables were included as covariates in the design matrix. Count data were normalized using the TMM method, incorporated into a limma object, and voom-transformed. Linear models were fitted for each gene, and contrasts were applied to identify differentially expressed genes for each group comparison.

#### Pathway Enrichment Analysis

Pathway enrichment was performed on the differentially expressed genes using clusterProfiler(v4.10.1). A filter of 0.05 was applied to the adjusted p-values (method = “BH”). The enrichment analysis was performed for upregulated (logFC > 0) and downregulated (logFC < 0) genes for the GO pathway databases using the function enrichGO(). A data frame containing term ID, pathway or category description, GeneRatio, and adjusted p-value was used.

To visualize gene- and pathway-level expression trends, expression data were normalized and batch-corrected using Surrogate Variable Analysis (SVA). For each pathway (lysosomal function, mitochondrial, ubiquitin–proteasome, interferon signaling, organelle stress, metabolism, phagocytosis), mean expression was calculated per group and center-scaled within pathways. Scaled values were plotted as a heatmap, enabling comparison of relative up- or downregulation across groups while minimizing inter-pathway variability.

#### Weighted Gene Co-Expression Analysis (WGCNA)

Weighted gene co-expression network analysis (WGCNA) was applied to corrected gene expression data in order to construct gene correlation networks and co-expression modules using the WGCNA(v1.72-5) package^94^. The counts were corrected using the same parameters as in the differential analysis for consistency. Using the sva_network package, we computed the SV loadings of the standardized expression matrix with singular value decomposition (SVD) and computed the residuals after regressing the top 12 SVs. Linear regression between the SVs and the covariates showed correlation mostly with technical covariates, including lane, batch, percentage of ribosomal bases and other sequencing metrics such as % of mRNA and intergenic bases (**Supplementary** Fig. 4A).

The co-expression network analysis was performed using the R package of Weighted Gene Correlation Network Analysis (WGCNA)^94^ following the standard pipeline to fit a scale-free topology (R2 > 0.8) and applying a Soft Threshold power of 5 into a signed network model (**Supplementary** Fig. 4B). The adjacency matrices were constructed using the average linkage hierarchical clustering of the topological overlap dissimilarity matrix (1-TOM). Co-expression modules were defined using a dynamic tree cut method with minimum module size of 20 genes and deep split parameter of 4. Modules highly correlated with each other, corresponding to a module eigengene (ME) correlation of 0.75 were merged, (**Supplementary** Fig. 4D). The genes were prioritized based on their module membership value, also known as eigengene-based connectivity (kME). The genes for each module and their gene-set enrichment analysis are shown in Supplementary Table 2. A Wilcoxon rank sum test was performed across modules comparing GBA1/iPD, LRRK2/iPD, GBA1/LRRK2, and iPD/Control groups to determine significance. The parameters used are paired is false (two-sided, Wilcoxon rank-sum test) and the p-adjust.method uses the Benjamini-Hochberg (BH) method. The significance criteria used was filtering at an adjusted p value less than 0.05.

To assess the enrichment of WGCNA modules within differentially expressed genes (**Fig. 2A-II**), and enrichment of custom gene-sets within modules (**Fig. 2A-III**), we performed Fisher’s exact tests to calculate enrichment ratios comparing the observed overlap of module genes with up- or down-regulated genes in GBA-PD and LRRK2-PD versus iPD or enrichment of custom gene-sets within module genes.

#### Visualization and Plots

All plots were created using ggplot2(version 3.5.0) in R(v4.3.3), with ggrepel(v0.9.5), and ggfortify(v0.4.17) for additional layers of visualization.

## Code availability

The code used for the primary analysis is available on GitHub at: https://github.com/RajLabMSSM/GBA-LRRK2-monocytes. Short-read RNA-seq pipeline: https://github.com/CommonMindConsortium/RAPiD-nf/. Any additional code used for analysis is available upon request from the corresponding author.

## Data Availability

All summary statistics are provided in the Supplementary Tables. Raw RNA-seq data and processed read counts will be made available via GEO upon publication. Raw RNA-seq data and genotype information for a subset of samples will also be deposited in dbGaP under accession ID phs002400.v1.p1.

https://github.com/RajLabMSSM/GBA-LRRK2-monocytes

https://github.com/CommonMindConsortium/RAPiD-nf/

## Acknowledgments

We thank the study participants for their blood donations to this study. T.R. is supported by grants from the U.S. National Institutes of Health (NIH) grants, including NINDS R01-NS116006, R01-R01-NS133742, NINDS U01-NS107016, NIA R21-AG063130, NIA R01-AG054005, NIA U01-AG068880, NIA RF1-AG065926, NIA R56-AG055824, NIA P30-AG066514, and NINDS U54-NS123743 and. R.S-P. is supported by NIH NINDS U01-NS107016, U01-NS094148-01, the Bonnie and Tom Strauss Chair, and the Bigglesworth Family Foundation. M.R. is supported by NIH F31-NS134319-01. We acknowledge contributions from Oriol Narcis Majos (O.N.M.) and Daniele Mattei (D.M.) and Casey Young (C.Y.) who are supported by the Silverstein Foundation Fellowship. Additional support was provided by the NIH Office of Research Infrastructure under award numbers S10OD018522 and S10OD026880.

This work was supported in part through the computational resources and staff expertise provided by Scientific Computing at the Icahn School of Medicine at Mount Sinai and supported by the Clinical and Translational Science Awards (CTSA) grant UL1TR004419 from the National Center for Advancing Translational Sciences. Research reported in this paper was supported by the Office of Research Infrastructure of the National Institutes of Health under award number S10OD026880 and S10OD030463. The content is solely the responsibility of the authors and does not necessarily represent the official views of the National Institutes of Health.

## Author Contributions

T.R. and R.S.P conceived and designed the study. D.M., O.N.M., E.M. and T.K. processed samples and generated the data. E.B., B.M. and D.M. analyzed the data and performed statistical analyses with assistance from M.R., and J.H., supervised by T.R.. D.M, A.D. RSP and T.R. interpreted the results. D.M. and T.R. wrote the manuscript. All the authors have read and approved the final manuscript. T.R. supervised the study.

