## Supplementary Figures for "Integrated Stress Response Signatures Drive Monocyte Dysfunction in *GBA1*- and *LRRK2*-Linked Parkinson’s Disease"

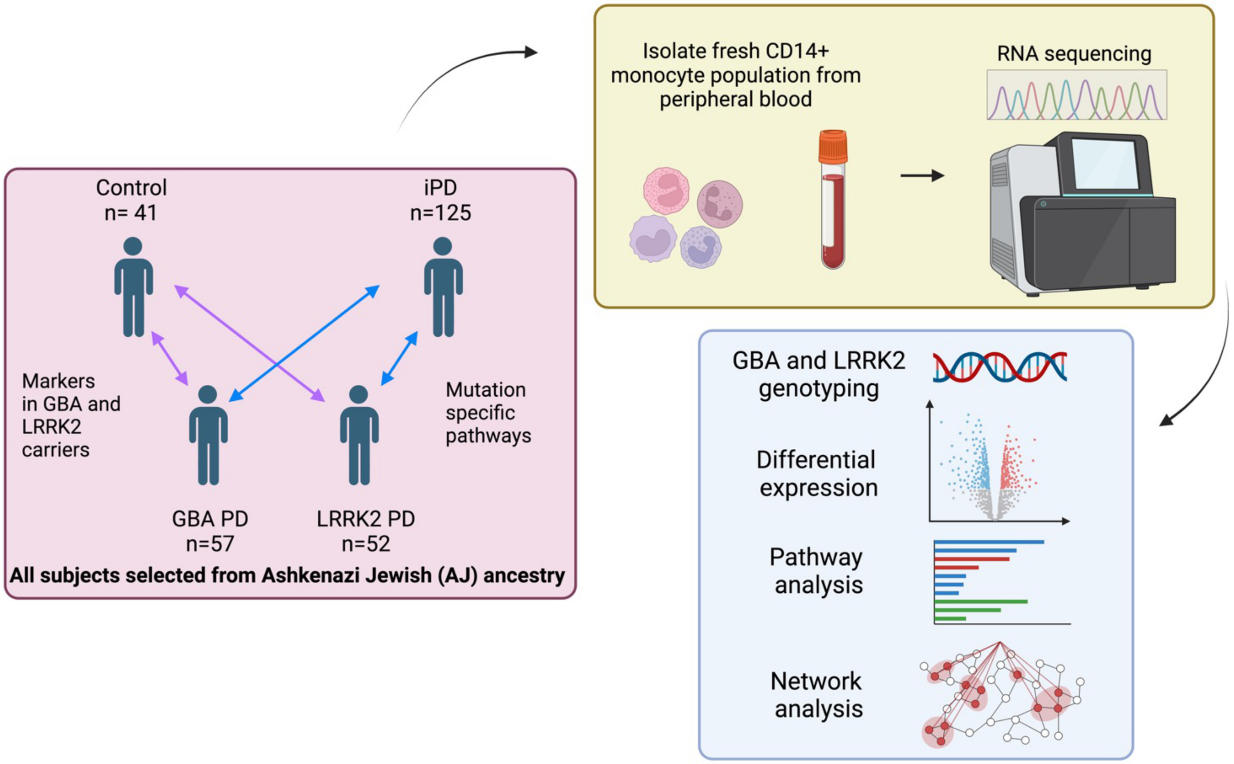


**Supplementary Fig. 1 | Project design schematic representation.** Schematic representation of project design and rationale for the comparison of the selected cohorts and analysis of biological samples in monocytes. Fresh CD14+ monocytes were isolated from peripheral blood within 3 hours of blood drawn from 233 donors with neurological diseases, as well as 41 unaffected subjects (controls) generating a total of 274 samples. Genome-wide genotyping was performed using DNA isolated from all donors and all subjects are from Ashkenazi Jewish European ancestry. The following analyses were performed with this dataset: (i) differential expression analysis; (ii) weighted gene co-expression analysis; (iii) gene set enrichment analysis.

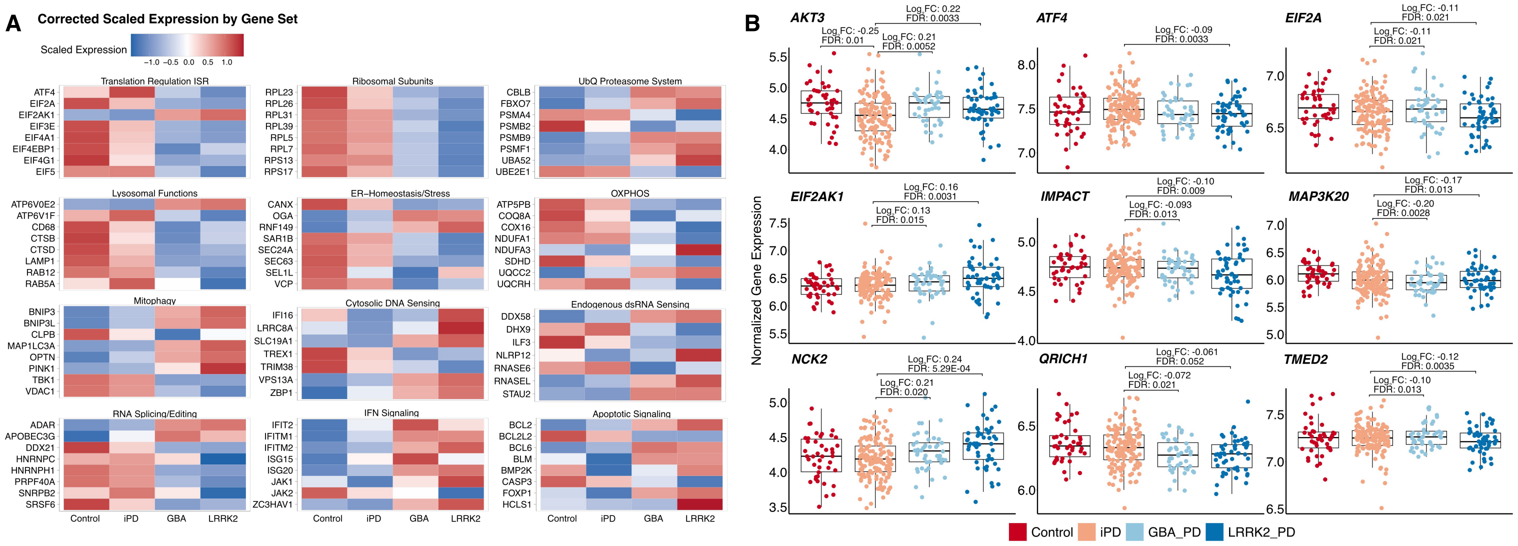


**Supplementary Fig. 2 | (A)** Heatmaps comparing the corrected, scaled SVA expression key genes associated with the biological processes significantly deregulated in GBA1- and LRRK2-PD monocytes relative to iPD (resolved from Fig. 1F). All displayed genes were significantly differentially expressed in either or both GBA1- and LRRK2-PD monocytes (FDR < 0.05, method = BH). Scaled expression is defined as z-score-transformed mean expression per gene, calculated by centering and standardizing SVA-corrected normalized expression. **(B)** Boxplots comparing the SVA corrected expression of core genes involved in the integrated stress response (ISR) signaling. Displayed genes are from the gene ontology term GO:0140467 (ISR signaling).

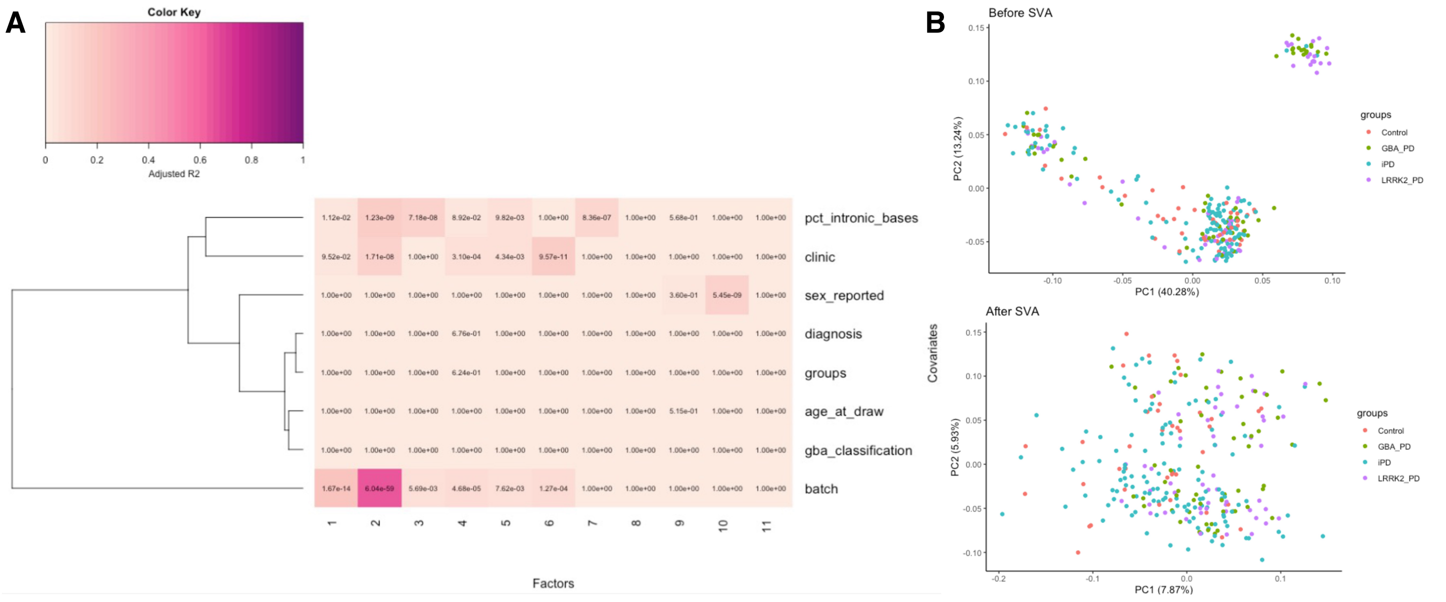


**Supplementary Fig. 3 | Covariate selection and correlation with possible confounders. (A)** Heatmap showing the correlation of the first 11 SVs (x-axis) and the known covariates (y-axis). (**B**) Scatter plot showing the results from the principal component (PC) analysis before and after surrogate variable analysis (SVA).

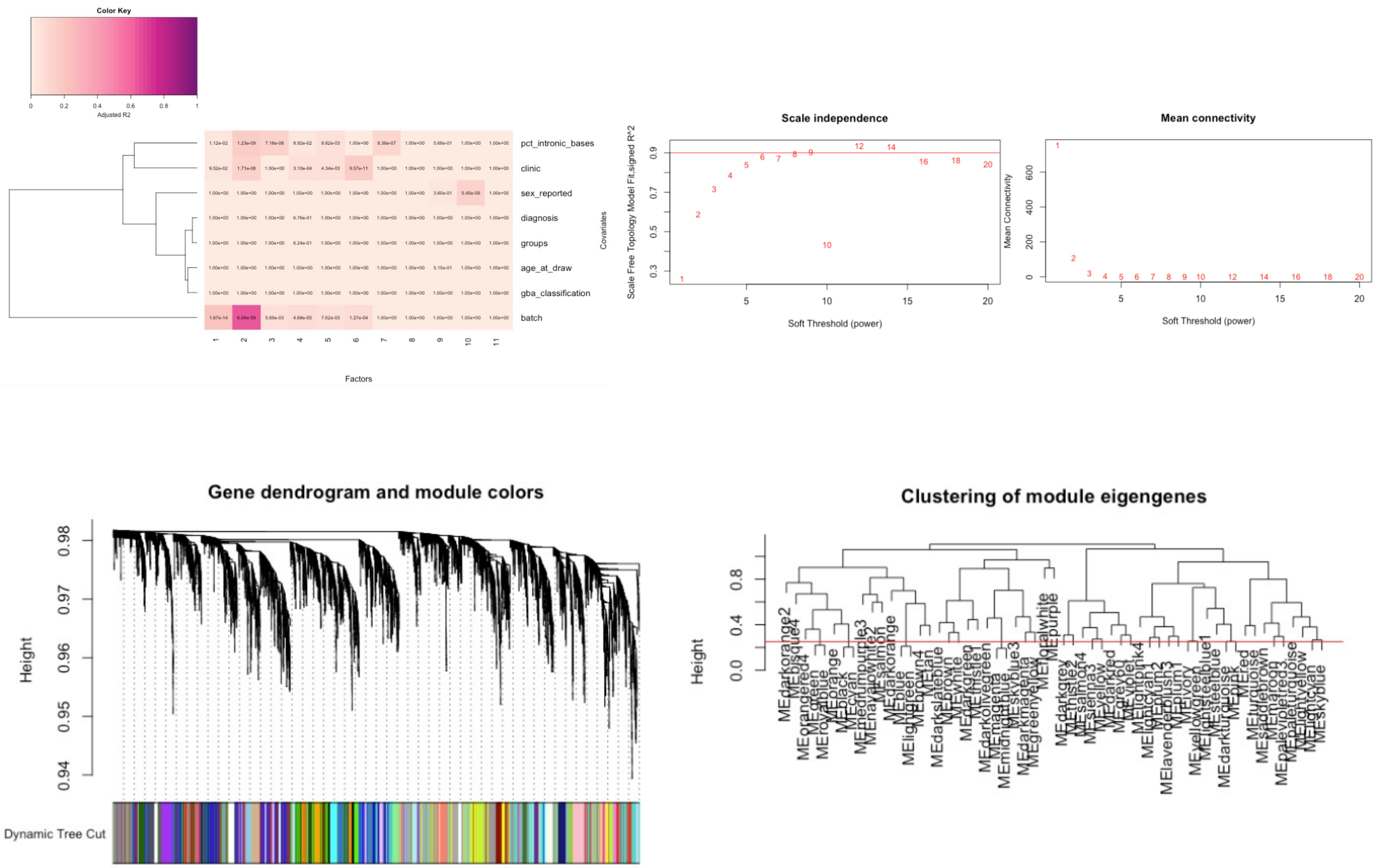


**Supplementary Fig. 4 | Gene network construction in human monocytes using WGCNA.** 53 unique co-expression modules were obtained using WGCNA. **(A)** Heatmap showing the correlation of the first 11 SVs (x-axis) and the known covariates (y-axis). **(B)** Right: Evaluation of network topology with different soft-thresholding powers. The y-axis represents the scale-free fit index as a function of the soft-thresholding power (x-axis). Left: The mean connectivity (y-axis) as a function of the soft-thresholding power. **(C)** Gene dendrogram using "Dynamic Tree Cut" to assign genes to different modules and modules to colors, showing before and after collapsing modules into 65 final networks. **(D)** Module eigengenes clustering dendrogram based on topological overlap. Modules below the threshold (Module Dissimilarity = 0.25) indicated by the red line were merged. These values correspond to a correlation of 0.75.

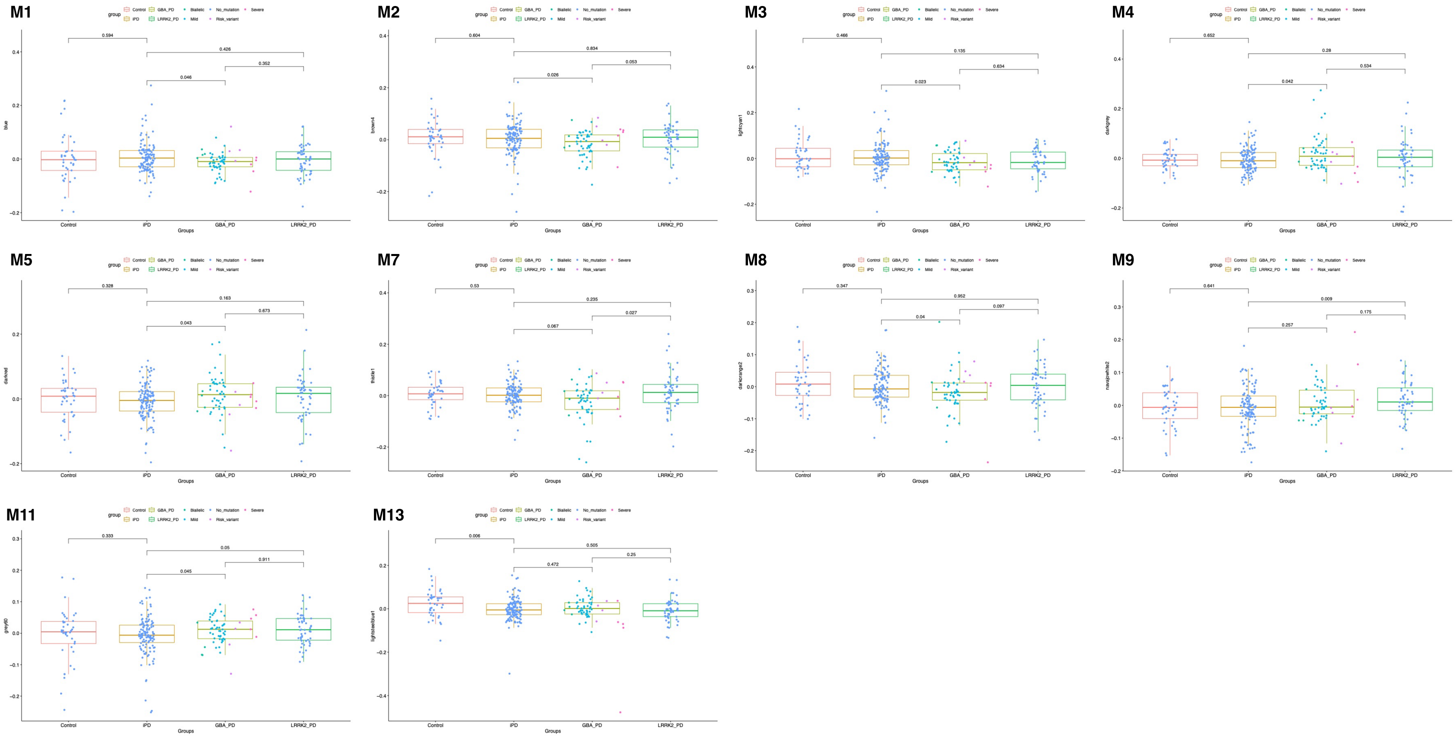


**Supplementary Fig. 5 | Modules Eigengene expression comparison between groups.** Boxplots show eigengene expressions for all modules except M6, M10 and M12, displayed in Fig. 3B. Boxplots indicate median, quartiles, and whiskers (1.5× IQR). Eigengene expression was compared via Wilkoxon sum-rank test, nominal p-value thresholds: *p ≤ 0.05, **p ≤ 0.01, *p ≤ 0.001.
